# Effectiveness of the Risk Assessment and Management Programme for hypertension (RAMP-HT) in reducing complications and service utilization after 5 years: A population-based matched cohort study

**DOI:** 10.1101/2022.09.12.22279844

**Authors:** Esther YT Yu, Eric YF Wan, Ivy L Mak, David VK Chao, Welchie WK Ko, Maria Leung, Yim Chu Li, Jun Liang, Luk Wan, Michelle MY Wong, Ruby LP Kwok, Anca KC Chan, Daniel YT Fong, Cindy LK Lam

**Affiliations:** Department of Family Medicine and Primary Care, School of Clinical Medicine, Li Ka Shing Faculty of Medicine, The University of Hong Kong; Department of Family Medicine and Primary Care, The University of Hong Kong – ShenZhen Hospital; Centre for Safe Medication Practice and Research, Department of Pharmacology and Pharmacy, Li Ka Shing Faculty of Medicine, the University of Hong Kong, Hong Kong; Laboratory of Data Discovery for Health (D24H), Hong Kong Science and Technology Park, Sha Tin, Hong Kong; Department of Family Medicine, Kowloon East Cluster, Hong Kong Hospital Authority, Hong Kong; Department of Family Medicine, Hong Kong West Cluster, Hong Kong Hospital Authority, Hong Kong; Department of Family Medicine, New Territories East Cluster, Hong Kong Hospital Authority, Hong Kong; Department of Family Medicine, Kowloon Central Cluster, Hong Kong Hospital Authority, Hong Kong; Department of Family Medicine, New Territories West Cluster, Hong Kong Hospital Authority, Hong Kong; Department of Family Medicine, Kowloon West Cluster, Hong Kong Hospital Authority, Hong Kong; Department of Family Medicine, Hong Kong East Cluster, Hong Kong Hospital Authority, Hong Kong; Hospital Authority Head Office, Hong Kong Hospital Authority, Hong Kong; New Territories East Cluster Statistics Office, Prince of Wales Hospital, Hong Kong Hospital Authority, Hong Kong; School of Nursing, Li Ka Shing Faculty of Medicine, The University of Hong Kong

**Keywords:** cardiovascular disease, health service utilization, hypertension, mortality, primary health care, risk-stratified management, team-based care

## Abstract

**Background:** The Risk Assessment and Management Programme for Hypertension (RAMP-HT) is a multi-component team-based intervention implemented in public primary care clinics across Hong Kong since 2011. After 12 months, significantly greater proportion of RAMP-HT participants achieved target blood pressure (Odd Ratio (OR) 1.18, P<0.001) and low-density lipoprotein-Cholesterol levels (OR 1.13, P<0.001) compared to usual care patients. This study evaluated the effectiveness of RAMP-HT for reducing hypertension-related complications and health service utilization among patients with hypertension relative to usual care after 5 years.

**Methods and Findings:** *Design:* Population-based prospective matched cohort study

*Setting:* All 73 public primary care clinics in Hong Kong

*Participants:* 212,707 adults with uncomplicated hypertension managed in public primary care clinics in Hong Kong between 2011 and 2013 were included. 108,045 RAMP-HT participants were matched to 104,662 patients receiving usual care using propensity score fine stratification weightings.

*Main outcome measures:* Hypertension-related complications (cardiovascular diseases (CVD), end-stage renal disease (ESRD)), all-cause mortality, public health service utilization (overnight hospitalization, attendances at accident and emergency department (AED), specialist outpatient clinic (SOPC) and general outpatient clinic (GOPC).

*Results:* After a median follow-up of 5.4 years, RAMP-HT participants had 8.0%, 1.6% and 10.0% lower absolute risks for CVD events, ESRD and all-cause mortality, respectively, compared to patients receiving usual care. After adjusting for all baseline covariates, the RAMP-HT group was associated with a 38% (Hazard Ratio (HR) 0.62, (95% CI 0.61 to 0.64)), 46% (HR 0.54, (95% CI 0.50 to 0.59)), and 48% (HR 0.52, (95% CI 0.50 to 0.54)) lower risk of CVD, ESRD, and all-cause mortality respectively, compared to the usual care group. The number-needed-to-treat was 16 to prevent one CVD event, 106 for ESRD and 17 for all-cause mortality. RAMP-HT participants also had significantly lower incidences for overnight hospitalization, AED, and SOPC attendances (Incidence Rate Ratio (IRR) 0.60, 0.70 and 0.87, respectively) but more GOPC attendances (IRR 1.06) compared to usual care patients.

**Conclusions:** RAMP-HT was associated with significant and clinically important reductions in all-cause mortality and hypertension-related complications for patients with hypertension in the naturalistic primary care setting after 5 years.

**Trial registration:** **NCT02219958**

## Introduction

Hypertension is the leading global risk factor for mortality and morbidity resulting from coronary heart diseases (CHD), stroke and end-stage renal disease (ESRD).[1] However, only one in three patients receiving treatment for hypertension worldwide had achieved blood pressure (BP) control.[2] Barriers to BP control include the lack of comprehensive primary care services to effectively deliver prevention and treatment at the system level,[3] poor adherence to clinical guidelines at the provider level, and noncompliance to medication and lifestyle intervention at the patient level.[4, 5] A multitude of implementation strategies, including team-based care, standardized management protocols, provider-training and patient empowerment, were found to be effective in reducing patients’ BP.[6-8] Multi-level, multi-component strategies were suggested to be the most effective for systolic BP (SBP) reduction.[9] However, the follow-up periods for these trials were relatively short (<24 months). Evidence for longer-term effects on cardiovascular outcomes, mortality, or health service utilization remains sparse. Furthermore, the impact of implementing these interventions on the population level is unknown.

Hong Kong has a dual health care system for its 7.5 million population, with a universal-type public health care system providing over 90% of hospital-based services, 70% of chronic disease care and approximately 30% of acute episodic care.[10] The public system is strained by the ever-rising service demand due to an aging population, increasing prevalence of chronic diseases and limited manpower.[11] On average, each public outpatient clinic consultation lasts for 3-5 minutes.[12] During a primary care consultation, each patient presents with an average of 1.3 health problems that may include both new complaints and chronic diseases.[13] Consequently, limited time can be spent on chronic disease care, contributing to therapeutic inertia among clinicians, poor patient-provider communication, and patients’ nonadherence to treatment due to the paucity of education and structured support. As of 2010, more than 45% patients with hypertension receiving treatment at public primary care clinics in Hong Kong had uncontrolled BP according to an internal report.

To improve the quality of hypertension care at the public primary care setting, the territory-wide Risk Assessment and Management Programme for hypertension (RAMP-HT) was launched to augment usual care since 2011 by the Hospital Authority (HA), the public health service provider in Hong Kong.[14] The RAMP-HT is a multi-level, multi-component, team-based intervention targeting at total cardiovascular disease (CVD) risk control of primary care patients with hypertension. The programme incorporates 3 main strategies: 1) task shifting from doctor to nurses for CVD risk assessment, education and empowerment of patients, and to allied health professionals for lifestyle interventions; 2) provider training with a standardized protocol; and 3) clinical decision support for team members using an enhanced electronic information-rely system.

Preliminary results from the programme showed that RAMP-HT participants who had suboptimal BP at baseline were significantly more likely to achieve target BP (odds ratio (OR) 1.18) and low-density lipoprotein-cholesterol (LDL-C) levels (OR 1.13) compared to patients receiving usual care after 12 months.[15] Notably, the observed differences between the two groups in SBP control (−0.84mmHg, P<0.01), LDL-C level (−0.05mmol/L, P<0.01) and predicted 10-year CVD risk (−0.39%, P<0.01) were only modest even though statistically significant. It remains uncertain whether the modest short-term reduction of these surrogate CVD markers can be translated to reduction of morbidity and mortality over time, and whether RAMP-HT is beneficial for all patients with hypertension, including those having controlled BP but with other uncontrolled CVD risk factors, in the naturalistic primary care setting.

## Methods

### Study Designs

This is a territory-wide prospective cohort study that compares the risks of CVD, ESRD, all-cause mortality, and frequencies of public health service utilization over five years between RAMP-HT participants and patients with hypertension who are receiving usual public primary care in Hong Kong.

### Settings

#### Usual care

Patients with hypertension are scheduled to attend a doctor consultation at a primary care clinic (i.e. general outpatient clinic (GOPC)) every 8 to 16 weeks. During a typical 5-minutes consultation, the attending primary care doctor may review the patient’s BP and control of other risk factors, titrate medication, advise on lifestyle, arrange assessment or refer to allied health professionals as appropriate, according to the Hong Kong reference framework for hypertension in primary care.[16] Based on clinical judgement, the doctor may also focus on managing the patient’s other health problems not directly related to hypertension care during the follow-up appointment. The patients are not required to register with a specific clinic or a regular doctor.

#### RAMP-HT

The RAMP-HT introduced three specific services to augment the time-constrained usual care: risk assessment, nurse intervention and specialist consultation.

All participants will undergo a risk assessment conducted by a trained nurse, which involves standardized CVD risk assessment (including smoking habit, BP level, lipid profile, glycaemic status and obesity), hypertensive complication screening (including the presence of left ventricular hypertrophy, arrhythmia, ischaemic heart disease, cerebrovascular disease, peripheral vascular disease, chronic kidney disease and proteinuria), and detailed assessment on self-care (including medication adherence, lifestyle and self-BP monitoring practice). Each participant is then stratified as having low, medium or high risk according to the 10-year predicted CVD risk calculated using the Joint British Societies’ (JBS2) risk calculator.[17] Based on individual risk level, presence of other CVD risk factors and participants’ preference, the nurse, serving as care-manager, empowers the participants on lifestyle and self-BP monitoring, prepares a care plan and coordinates follow-up interventions by a multi-disciplinary team for the participants according to protocol (**S1 Fig**). To enhance communication among team members and facilitate clinical decision-making of team members, the care-manager records each participant’s CVD risk profile and care plan on an electronic record platform with an action reminder system. The risk assessment may be repeated every 12-30 months depending on the participant’s risk profile.

Participants with adherence issues or specific risk factors are referred for nurse interventions; participants with resistant hypertension (i.e. SBP ≥ 160/100 mm Hg despite taking at least 3 anti-hypertensive medications) are referred for additional specialist consultations. Meanwhile, all participants continue to attend their usual primary care doctors every 8-16 weeks at GOPCs, who receive an electronic report on the participants’ CVD risk control and additional intervention received, and an action reminder to facilitate medication titration. The usual care doctors may also provide care for other co-morbidities and new health problems. Availability of medications, access to laboratory tests, investigations and allied health professional services in the public primary care clinics and community centers are the same for RAMP-HT participants and usual care patients.

To implement the RAMP-HT as an add-on intervention to usual care, RAMP-HT services were gradually set up at 59 out of 73 GOPCs in Hong Kong between 2011 to 2013 to serve patients residing in different geographic clusters. All patients aged 18 years or above who had hypertension and were receiving management in any of the 73 GOPCs, but no pre-existing diabetes mellitus (DM), CVD or ESRD were eligible to participate in the RAMP-HT, while patients with concomitant DM were channeled for enrollment in the parallel Risk Assessment and Management Programme for Diabetes Mellitus (RAMP-DM) instead.[18] Two cycles of evaluation on the quality of care of the RAMP-HT were conducted in 2013 and 2015, which confirmed satisfactory adherence to the RAMP-HT protocol. Although the RAMP-HT is a territory-wide programme intended for all patients with hypertension managed in GOPCs, roll-out of the programme could only be performed in stages due to the large number of patients, limited resources and manpower. Patients were randomly invited by their attending usual care doctors to voluntarily enroll in the RAMP-HT during their scheduled follow-up visits. Consequently, there was a small opportunity window to compare the effect of RAMP-HT and usual care in the real-world primary care setting.

### Study Participants

The inclusion criteria for this study are: 1) aged 18 years or above; 2) diagnosed with hypertension – defined by the International Classification of Primary Care-2 (ICPC-2) code of K86 on or before baseline; 3) not diagnosed with DM – defined by ICPC-2 codes of T89 or T90 before baseline; 4) without a CVD or ESRD diagnosis (defined below) based on historical medical records, and; 5) received hypertension care from one of the 73 GOPCs. Electronic health records of eligible patients were extracted from the HA Clinical Management System. RAMP-HT participants consisted of patients who enrolled in RAMP-HT between 1^st^ October 2011 and 30^th^ September 2013; usual care patients were those who had visited any GOPC at least once for hypertension care within the same period, but had not been enrolled in the RAMP-HT by 30^th^ September 2017. The baseline dates for the RAMP-HT and usual care groups were defined as the first date of attending a risk assessment session and the first date of attending GOPC during the aforementioned period, respectively. For each outcome (defined below), each patient was observed from his/her baseline date to the date of all-cause mortality, incidence of an outcome event or last follow-up censored until 30^th^ September 2017, whichever occurred first.

### Outcome Measures

The primary composite outcome was any CVD events, including CHD, heart failure or stroke; ESRD; or all-cause mortality. Secondary outcomes included: 1) CVD; 2) CHD; 3) stroke; 4) heart failure; 5) ESRD; 6) DM; 7) all-cause mortality; and 8) public health service utilization, including overnight hospitalization, and attendances at accident & emergency department (AED), specialist outpatient clinic (SOPC) and GOPC. The definitions of events were described in **S1 Table**. Mortality data were extracted from the Hong Kong Death Registry.

### Baseline Covariates

Baseline covariates comprised of socio-demographics, medical history, anthropometric measurements and laboratory results of patients. Socio-demographics included gender, age and smoking status. Medical history included the Charlson Comorbidity Index, prescriptions of anti-hypertensive medications including angiotensin converting enzyme inhibitor/angiotensin receptor blocker (ACEI/ARB), β-blocker, calcium channel blocker (CCB), diuretic or other anti-hypertensive medications, and lipid-lowering agents such as statins and fibrate. Anthropometric measurements included SBP and diastolic BP (DBP) measured at GOPC using semi-automated blood pressure measurement devices, and body mass index (BMI). Laboratory results consisted of full lipid profile (LDL-C level, total cholesterol to high-density lipoprotein-cholesterol ratio (TC/HDL-C ratio) and triglyceride level), fasting glucose (FG) level, and estimated glomerular filtration rate (eGFR) calculated based on creatinine level from blood test according to the abbreviated Modification of Diet in Renal Disease Study formula recalibrated for Chinese.[19] All laboratory assays were performed in laboratories accredited by the College of American Pathologists, the Hong Kong Accreditation Service or the National Association of Testing Authorities, Australia.

### Data Analysis

Missing baseline covariates were handled by multiple imputation. Specifically, missing values were imputed five times by the chained equation method using all known baseline covariates and event outcomes.[20] The same analysis was performed for each dataset and the five sets of results were combined using Rubin’s rules.[21]

To reduce potential selection bias, all RAMP-HT participants and usual care patients were matched by propensity score fine stratification weightings, which is an extension of propensity score matching that combines propensity score stratification with weighting technique.[22] The propensity score for each patient was generated by fitting a logistic regression with the patient’s corresponding RAMP-HT or usual care group as a dependent variable and all baseline covariates as independent variables. To produce comparable baseline characteristics between groups, fine stratification weightings assigned weights to each patient based on the stratified propensity score. Fine stratification weightings were generated by the ‘MMWS’ package in Stata with thousand quintile categories of propensity scores for each stratum.[23] After weighting, balance of baseline covariates between the two groups was further assessed using standardized mean difference (SMD); SMD of less than 0.2 implies sufficient balance between the groups.[24]

Summary statistics were described in mean (standard deviation) or frequency (proportion). Differences in baseline characteristics were compared using univariable linear, logistic, multinomial logistic or negative binomial regression, as appropriate. 5-year cumulative incidences, incidence rates of each outcome and service utilization, and corresponding absolute risk reduction (ARR) in each group were reported. Kaplan-Meier survival curves for incidences of event outcomes between groups were compared by the log-rank test. Multivariable Cox proportional regressions were performed to estimate the effect of RAMP-HT on the risk of each event outcome adjusted for baseline characteristics. Proportional hazards assumption was checked by examining plots of the scaled Schoenfeld residuals against time for the covariates. Multi-collinearity was assessed by the variance inflation factor. All models satisfied the proportional hazards assumption and no multi-collinearity existed after assessing the variance inflation factor. Number needed to treat (NNT) for each outcome was calculated based on corresponding hazard ratios (HR).[25] Frequencies of health service utilizations were compared using negative binomial regression adjusted for baseline characteristics and corresponding incidence rate ratios (IRRs) were calculated.

Six sensitivity analyses were conducted to evaluate the robustness of the results. First, to minimize reverse causality, matched patients with less than 1-year follow-up were excluded from the analysis. Second, one-to-one propensity score matching with multiple imputation but no fine stratification weighting was used to match the baseline characteristics of the patients. Third, all eligible RAMP-HT participants and usual care patients were included in the analysis after multiple imputation without propensity score matching or weightings. Fourth, fine stratification weightings without multiple imputation was performed. Fifth, propensity score matching without multiple imputation was conducted. Lastly, a complete case analysis that included only patients with complete datasets was performed.

The effects of RAMP-HT on all outcomes were also compared in subgroups, stratified by gender (male, female), age (<65years, ≥65years), smoking status (past- or non-smoker, smoker), SBP (<140mm Hg, 140-159mm Hg, ≥160mm Hg), fasting glucose (<6.1mmol/L, ≥6.1mmol/L), LDL-C (<3.4 mmol/L for Framingham 10-year CVD risk ≤20%; or <2.6 mmol/L for Framingham 10-year CVD risk >20%; other), eGFR (<60ml/min/1.73m^2^, ≥60ml/min/1.73m^2^), BMI (normal <23kg/m^2^, overweight 23-24.9kg/m^2^, obesity 25-30kg/m^2^, obesity class II ≥30kg/m^2^), estimated Framingham 10-year CVD risk (<10%, 10-19%, ≥20%), the Charlson Comorbidity Index (< 3, ≥ 3), number of anti-hypertension drugs prescribed (0, 1, 2, 3), classes of anti-hypertensive drug prescribed (ACEI/ARB, β-blocker, CCB, diuretic, other anti-hypertensive drugs), and lipid-lowering agents prescribed (statins, no statins) at baseline. Interactions between the RAMP-HT effect and each group were assessed, the significance of interactions were based on Hommel-adjusted P values.[26]

All tests were two-tailed and a p-value of less than 0.05 was considered statistically significant. Statistical analyses were performed in Stata Version 13.0.

### Ethics

The research protocol was approved by the Clinical and Research Ethics Committees of all 7 Hospital Authority geographical clusters in Hong Kong. Anonymous data were extracted through the HA Clinical Management System; consent from participants was not required.

## Results

A total of 212,707 primary care patients with hypertension (RAMP-HT n=108,045, Usual care n=104,662) met the inclusion criteria, were matched with fine stratification weightings and included in the analyses. The patient inclusion flow was illustrated in **S2 Fig**. Data completion rates for all baseline covariates ranged from 69.9% to 100% (**S2 Table**). Baseline characteristics of the RAMP-HT and usual care groups were similar before and after multiple imputations (**S3 Table**).

**Table 1** describes the characteristics of RAMP-HT participants (mean age 66.3 ± 12.3 years, 57.6% female) and usual care patients (mean age 66.3 ± 13.5 years, 57.8% female) at baseline after multiple imputations and fine stratification weightings. The SMD in all characteristics were below 0.2, indicating balance between the two groups. During the median follow-up period of 5.4 years, RAMP-HT participants received on average 2.25 risk assessments, 0.3 nurse intervention and 0.1 specialist consultation. At 5 years, a total of 72.9% and 67.3% RAMP-HT participants achieved target BP and LDL-C, respectively, compared to 66.5% and 61.8% in the usual care group (**Table 2)**. After adjustments for baseline values, RAMP-HT participants had 37% higher odds (OR 1.36, 95% CI 1.33 to 1.39) for achieving targets of all 5 CVD risk factors when compared to usual care patients. Characteristics of RAMP-HT and usual care patients at 5 years are shown in **S4 Table**. A greater proportion of RAMP-HT participants received statins prescription (increased from 7.8% at baseline to 39.4% at 5 years) compared to the usual care group (increased from 7.3% to 32.9%).

**Table 1.** Characteristics of RAMP-HT and usual care patients at baseline after multiple imputation and fine stratification weightings.

|  | RAMP-HT<br>(N = 108,045) | Usual Care<br>(N = 104,662) | SMD |
| --- | --- | --- | --- |
| **Socio-Demographic (% , n)** |  |  |  |
| Gender |  |  | 0.003 |
| Female | 57.6 % (62,277) | 57.8 % (60,497) |  |
| Age (mean±SD), year | 66.3±12.3 | 66.3±13.5 | <0.001 |
| Smoking Status |  |  | 0.007 |
| Smoker | 7.8 % (8,470) | 7.7 % (8,021) |  |
| **Clinical Characteristics (mean±SD)** |  |  |  |
| SBP, mm Hg | 136.6±16.5 | 136.5±17.8 | 0.004 |
| DBP, mm Hg | 76.3±11.4 | 76.3±11.6 | <0.001 |
| Fasting Glucose, mmol/L | 5.4±1.2 | 5.4±0.7 | <0.001 |
| LDL-C, mmol/L | 3.2±0.9 | 3.2±0.9 | 0.005 |
| TC/HDL-C Ratio | 4.0±2.2 | 4.0±1.6 | 0.001 |
| Triglyceride, mmol/L | 1.5±1.0 | 1.5±1.0 | <0.001 |
| BMI, kg/m<sup>2</sup> | 25.4±4.1 | 25.4±5.2 | <0.001 |
| eGFR (% , n) |  |  | 0.007 |
| ≥ 60ml/min/1.73m<sup>2</sup> | 95.2 % (102,906) | 95.4 % (99,849) |  |
| < 60ml/min/1.73m<sup>2</sup> | 4.8 % (5,139) | 4.6 % (4,813) |  |
| Charlson Comorbidity Index | 3.1±1.2 | 3.1±1.4 | 0.003 |
| Number of anti-hypertensive drugs prescribed (% , n) |  |  | 0.009 |
| 0-1 | 56.5 % (62,506) | 56.0 % (57,173) |  |
| ≥2 | 43.5 % (45,539) | 44.0 % (47,489) |  |
| Prescription of ACEI/ARB (% , n) | 19.7 % (21,259) | 19.4 % (20,253) | 0.008 |
| Prescription of β-blocker (% , n) | 37.0 % (40,026) | 37.0 % (38,763) | <0.001 |
| Prescription of CCB (% , n) | 70.4 % (76,077) | 70.3 % (73,571) | 0.003 |
| Prescription of Diuretic (% , n) | 12.5 % (13,521) | 12.5 % (13,041) | 0.002 |
| Prescription of other anti-hypertensive drugs (% , n) | 10.8 % (11,674) | 10.7 % (11,189) | 0.004 |
| Prescription of statin (% , n) | 7.8 % (8,447) | 7.3 % (7,685) | 0.018 |
| Prescription of fibrate (% , n) | 1.8 % (1,938) | 1.7 % (1,821) | 0.004 |
| **Frequency of Service Utilization† (mean±SD)** |  |  |  |
| Overnight Hospitalization | 0.2±1.1 | 0.2±0.5 | 0.007 |
| Accident & Emergency | 0.4±2.5 | 0.4±1.1 | 0.004 |
| Specialist Out-patient Clinic | 1.7±3.6 | 1.7±3.0 | 0.003 |
| General Out-patient Clinic | 5.6±3.5 | 5.6±2.7 | 0.023 |

RAMP-HT = Risk Assessment and Management Programme - Hypertension; SBP = Systolic Blood Pressure; DBP = Diastolic Blood Pressure; LDL-C = Low Density Lipoprotein - Cholesterol; TC = Total Cholesterol; HDL-C = High Density Lipoprotein - Cholesterol; BMI = Body Mass Index; eGFR = Estimated Glomerular Filtration Rate; ACEI = Angiotensin Converting Enzyme Inhibitor; ARB = Angiotensin Receptor Blocker; CCB = Calcium Channel Blocker; SD = Standard Deviation; SMD = Standardized Mean Difference
Notes:
† Service utilization was measured 1) at baseline: from a year before baseline to baseline; 2) at 5 years: from baseline to last follow-up.
SMD<0.2 indicates balance between groups.

The 5-year cumulative incidence of the primary composite outcome of any CVD or all-cause mortality was 11.8% in the RAMP-HT group and 26.3% in the usual care group (**Table 3**), corresponding to an absolute risk reduction of 14.5% for RAMP-HT participants. The differences in cumulative hazards for adverse outcomes between the two groups were apparent as early as 1-year after the programme (**Fig 1)**, and continued at least up to the fifth year. After adjusting for all baseline covariates, the RAMP-HT group was associated with a 42% (HR 0.58, (95% CI 0.57 to 0.59)) lower risk of composite outcomes compared to the usual care group. The NNT to prevent one composite event was 11. RAMP-HT participants had 38% (HR 0.62, (95% CI 0.61 to 0.64)), 46% (HR 0.54, (95% CI 0.50 to 0.59)), 17% (HR 0.83, (95% CI 0.80 to 0.85)) and 48% (HR 0.52, (95% CI 0.50 to 0.54)) lower risks of CVD, ESRD, DM and all-cause mortality respectively. The NNT to prevent one event was 16 for CVD, 106 for ESRD, 41 for DM and 17 for all-cause mortality. Consistently, hospital-based service utilizations including overnight hospitalization, AED and SOPC attendances were reduced by 40%, 30% and 13%, respectively in the RAMP-HT group (**Table 4**). The six sensitivity analyses showed similar results as the main analyses (**S5 and S6 Table**).

**Table 2.** Proportion of patients in the RAMP-HT or usual care achieving clinical targets at baseline and 5 years.

|  |  | Baseline | At 5 years | OR for RAMP-HT group achieving target adjusted for baseline value† |
| --- | --- | --- | --- | --- |
| SBP < 140 mm Hg & DBP < 90mm Hg | RAMP-HT Participants | 57.2% (57,330) | 72.9% (72,966) | 1.37 (1.34,1.40) * |
|  | Non-RAMP-HT Participants | 59.5% (50,178) | 66.5% (56,103) |  |
|  | OR for RAMP-HT achieving target | 0.94 (0.93,0.96) * | 1.35 (1.32,1.38) * |  |
| LDL-C (< 3.4 mmol/L for Framingham 10-year CVD risk ≤ 20%, or < 2.6 mmol/L for Framingham 10-year CVD risk > 20%) | RAMP-HT Participants | 44.0% (41,886) | 67.3% (64,012) | 1.31 (1.28,1.33) * |
|  | Non-RAMP-HT Participants | 44.6% (33,215) | 61.8% (46,048) |  |
|  | OR for RAMP-HT achieving target | 0.98 (0.97,1.00) | 1.27 (1.25,1.30) * |  |
| FG < 5.6mmol/L | RAMP-HT Participants | 68.5% (73,968) | 63.1% (60,966) | 1.22 (1.19,1.25) * |
|  | Non-RAMP-HT Participants | 67.6% (70,757) | 58.4% (46,170) |  |
|  | OR for RAMP-HT achieving target | 1.04 (1.02,1.06) * | 1.22 (1.20,1.24) * |  |
| BMI < 27.5 kg/m<sup>2</sup> | RAMP-HT Participants | 73.6% (79,566) | 72.9% (61,907) | 1.01 (0.98,1.05) |
|  | Non-RAMP-HT Participants | 72.8% (76,214) | 71.1% (42,451) |  |
|  | OR for RAMP-HT achieving target | 1.04 (1.02,1.07) * | 1.10 (1.07,1.12) * |  |
| Non-smokers | RAMP-HT Participants | 92.2% (99,575) | 93.6% (101,109) | 1.16 (1.10,1.24) * |
|  | Non-RAMP-HT Participants | 92.3% (96,641) | 93.4% (96,687) |  |
|  | OR for RAMP-HT achieving target | 0.98 (0.94,1.01) | 1.04 (1.00,1.08) |  |
| Achieved all 5 targets above | RAMP-HT Participants | 17.7% (19,132) | 58.7% (47,420) | 1.36 (1.33,1.39) * |
|  | Non-RAMP-HT Participants | 18.4% (19,252) | 51.8% (28,143) |  |
|  | OR for RAMP-HT achieving target | 0.95 (0.93,0.98) * | 1.32 (1.29,1.35) * |  |

RAMP-HT = Risk Assessment and Management Programme - Hypertension; SBP = Systolic Blood Pressure; DBP = Diastolic Blood Pressure; LDL-C = Low Density Lipoprotein - Cholesterol; FG = Fasting Glucose; BMI = Body Mass Index; CVD = Cardiovascular Disease; OR = Odds Ratio;
Notes:
\* Significant difference at 0.05 level by logistic regression with propensity score fine stratification weightings
† OR were adjusted for the corresponding target of clinical outcome at baseline

**Table 3.** Outcome events at 5 years in RAMP-HT participants and usual care patients.

| Event | RAMP-HT Participants (N = 108,045) |  |  | Usual Care Patients (N = 104,662) |  |  | ARR | HR† | P-value | NNT |
| --- | --- | --- | --- | --- | --- | --- | --- | --- | --- | --- |
|  | Cases with Event | Incidence | Incidence rate (Cases/100 Person Years) (95% CI) | Cases with Event | Incidence | Incidence rate (Cases/100 Person Years) (95% CI) |  |  |  |  |
| All outcome event | 12,784 | 11.8% | 3.1 (3.0,3.1) | 27,514 | 26.3% | 5.4 (5.3,5.5) | 14.5% | 0.58 (0.57,0.59) | <0.001* | 11 (10,11) |
| CVD | 9,167 | 8.5% | 2.1 (2.1,2.2) | 17,261 | 16.5% | 3.4 (3.3,3.4) | 8.0% | 0.62 (0.61,0.64) | <0.001* | 16 (15,16) |
| CHD | 3,964 | 3.7% | 0.9 (0.8,0.9) | 6,607 | 6.3% | 1.3 (1.2,1.3) | 2.6% | 0.66 (0.63,0.69) | <0.001* | 47 (44,51) |
| Heart Failure | 1,799 | 1.7% | 0.4 (0.4,0.5) | 5,094 | 4.9% | 0.9 (0.9,0.9) | 3.2% | 0.54 (0.51,0.58) | <0.001* | 49 (46,52) |
| Stroke | 4,378 | 4.1% | 1.0 (1.0,1.0) | 8,467 | 8.1% | 1.6 (1.5,1.6) | 4.0% | 0.64 (0.61,0.66) | <0.001* | 36 (33,38) |
| ESRD | 808 | 0.7% | 0.2 (0.2,0.2) | 2,409 | 2.3% | 0.4 (0.4,0.4) | 1.6% | 0.54 (0.50,0.59) | <0.001* | 106 (97,120) |
| DM | 10,235 | 9.5% | 2.4 (2.4,2.5) | 14,724 | 14.1% | 3.1 (3.1,3.2) | 4.6% | 0.83 (0.80,0.85) | <0.001* | 41 (36,48) |
| All-Cause Mortality | 4,833 | 4.5% | 1.2 (1.2,1.3) | 15,144 | 14.5% | 2.8 (2.7,2.8) | 10.0% | 0.52 (0.50,0.54) | <0.001* | 17 (16,18) |
| CVD Mortality | 1,532 | 1.4% | 0.4 (0.4,0.4) | 5,576 | 5.3% | 1.0 (1.0,1.1) | 3.9% | 0.51 (0.48,0.54) | <0.001* | 43 (40,46) |
| Non-CVD Mortality | 3,301 | 3.1% | 0.8 (0.8,0.8) | 9,568 | 9.1% | 1.8 (1.7,1.8) | 6.0% | 0.54 (0.51,0.56) | <0.001* | 45 (43,48) |

RAMP-HT = Risk Assessment and Management Programme - Hypertension; DM = Diabetes Mellitus; CVD = Cardiovascular Disease; CHD = Coronary Heart Disease; ESRD = End-Stage Renal Disease; CI = Confidence Interval; ARR = Absolute Risk Reduction; HR = Hazard Ratio; NNT = Number Needed to Treat
Notes:
\* Significant at 0.05 level by multivariable Cox proportional hazard regression with propensity score fine stratification weightings
† Hazard ratios were adjusted by gender, age, smoking status, SBP, DBP, fasting glucose, LDL-C, TC/HDL-C ratio, triglyceride, BMI, eGFR, Charlson Comorbidity Index and the usages of ACEI/ARB, β-blocker, CCB, Diuretic, other anti-hypertensive drugs, statin and fibrate at baseline

**Table 4.** Public health service utilization of RAMP-HT participants and usual care patients at 5 years.

| Service | RAMP-HT Participants (N = 108,045) |  |  |  |  | Usual Care Patients (N = 104,662) |  |  |  |  | IRR† (95% CI) | P-value |
| --- | --- | --- | --- | --- | --- | --- | --- | --- | --- | --- | --- | --- |
|  | Mean | Median | Range | Total Number of Events | Event Rate (Cases/100 Person Years) | Mean | Median | Range | Total Number of Events | Event Rate (Cases/100 Person Years) |  |  |
| Overnight Hospitalization | 1.11 | 0 | 0 - 69 | 119,670 | 23.4 | 1.90 | 1 | 0 - 105 | 199,951 | 36.7 | 0.60 (0.59,0.61) | <0.001* |
| A&E Attendance | 1.88 | 1 | 0 - 200 | 203,769 | 39.9 | 2.80 | 2 | 0 - 1073 | 293,703 | 53.9 | 0.70 (0.69,0.71) | <0.001* |
| SOPC Attendance | 9.05 | 4 | 0 - 126 | 979,227 | 191.5 | 10.86 | 6 | 0 - 254 | 1,141,245 | 209.6 | 0.87 (0.86,0.88) | <0.001* |
| GOPC Attendance | 21.03 | 20 | 0 - 422 | 2,276,471 | 445.2 | 22.27 | 22 | 0 - 1381 | 2,339,024 | 429.5 | 1.06 (1.06,1.06) | <0.001* |

RAMP-HT = Risk Assessment and Management Programme - Hypertension; A&E = Accident and Emergency; SOPC = Specialist Out-patient Clinics; GOPC = General Out-patient Clinics; IRR = Incidence Rate Ratio; CI = Confidence Interval
Notes:
\* Significant at 0.05 level by multivariable Cox proportional hazard regression/multivariable negative binomial regression with propensity score fine stratification weightings
† All Incidence rate ratios were adjusted by gender, age, smoking status, SBP, DBP, fasting glucose, LDL-C, TC/HDL-C ratio, triglyceride, BMI, eGFR, Charlson Comorbidity Index and the usages of ACEI/ARB, β-blocker, CCB, Diuretic, other anti-hypertensive drugs, statin and fibrate, and the corresponding number of service utilizations at baseline

**Fig 1.**
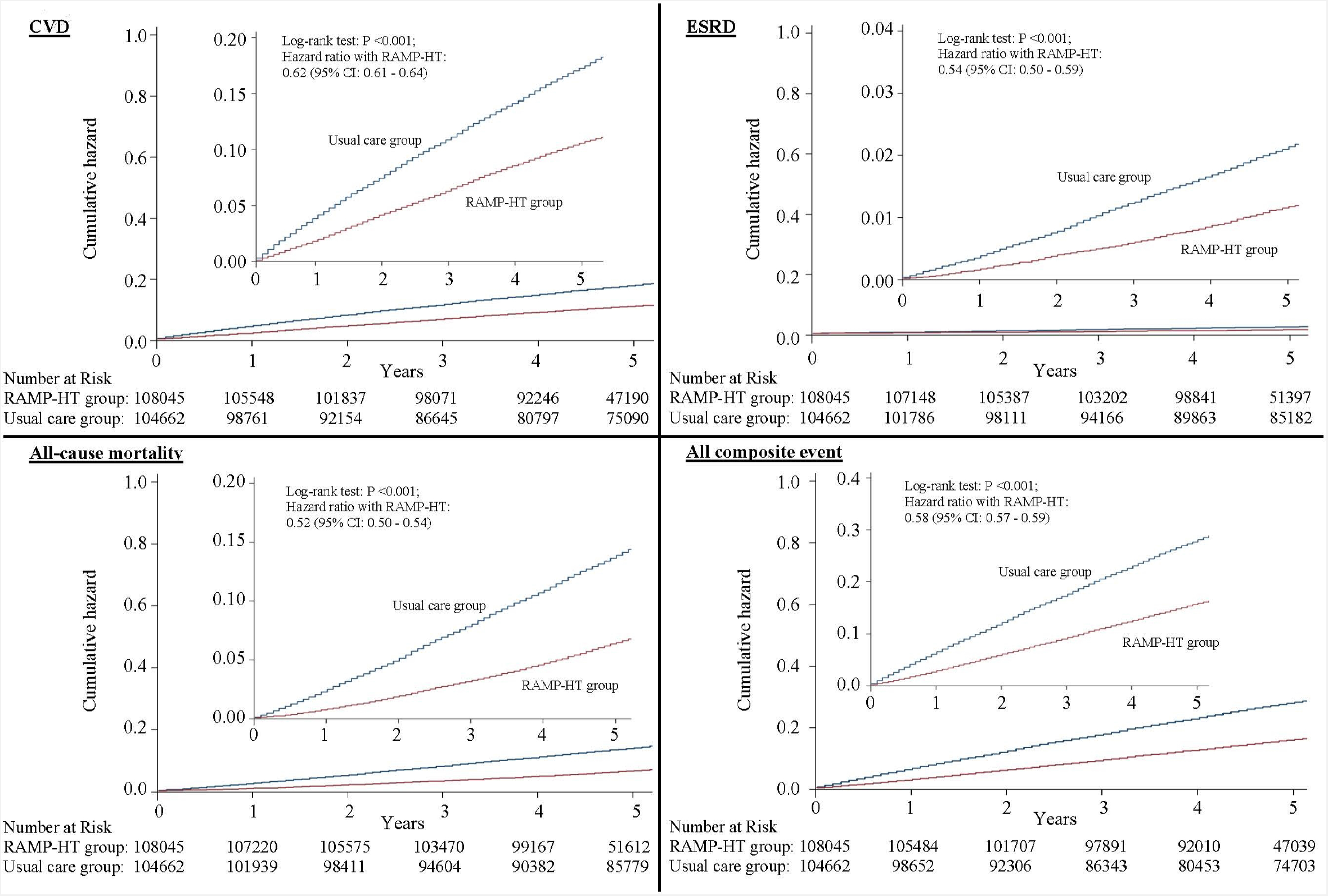
Cumulative hazards for cardiovascular disease (CVD), end-stage renal disease (ESRD), all-cause mortality and all composite event of primary outcome between RAMP-HT group and usual care group (CI = confidence interval)

Sub-group analyses were conducted to further determine the differential effects of participant characteristics on the incidences of adverse events (**Fig 2a**) and service utilization (**Fig 2b**). Similar to the main analyses, RAMP-HT participants had greater risk reduction in all event outcomes and lower hospital-based service utilization regardless of sub-groups. Only age and FG were found to have significant interactions with the effects of RAMP-HT. Participants < 80 years old showed 10% greater reduction in risks of adverse event outcomes compared to those who were older; similarly, the beneficial effects of RAMP-HT were approximately 13% greater among individuals with FG < 6.1mmol/L.

**Fig 2a.**
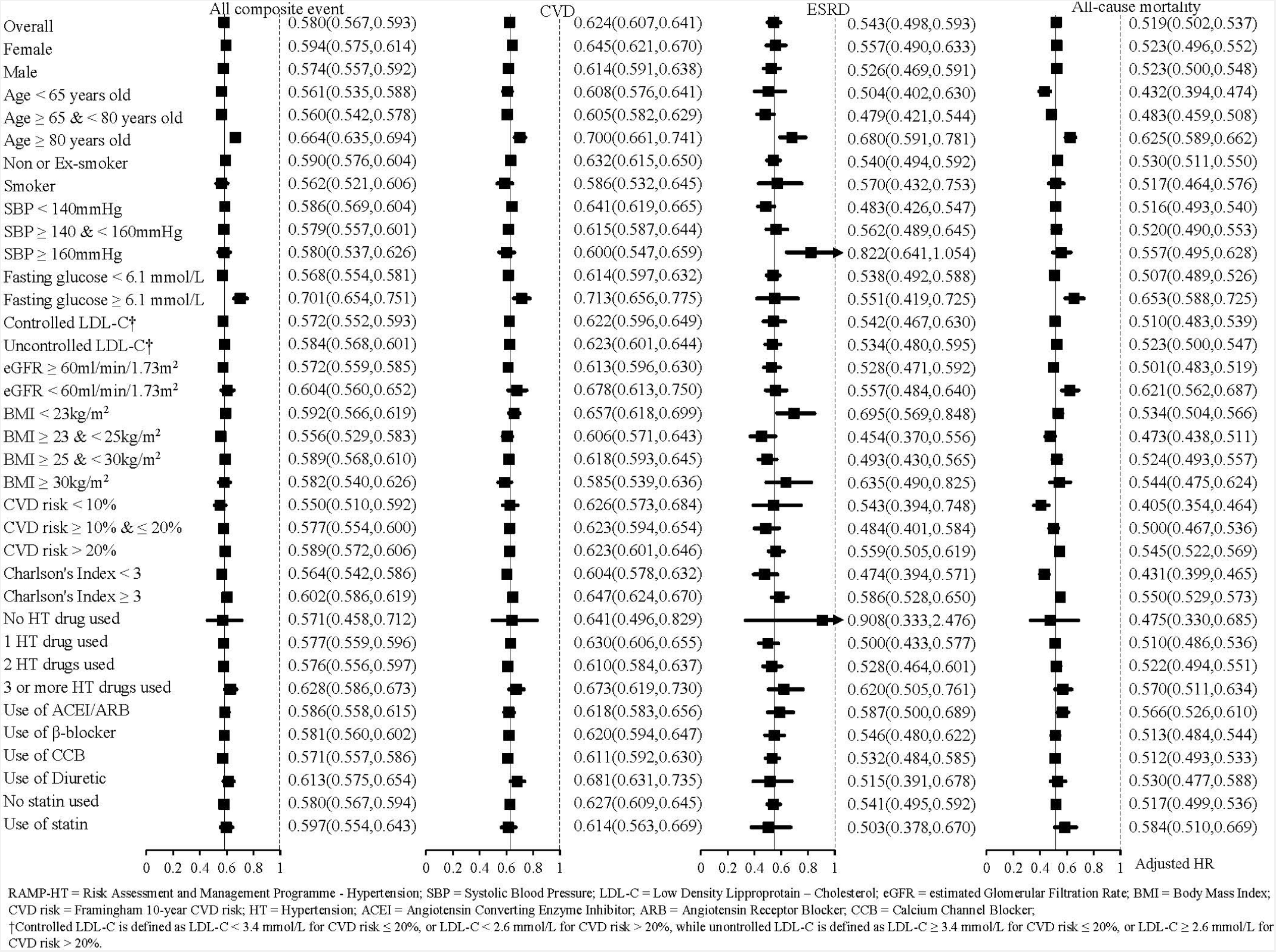
Adjusted hazard ratios (HRs) of RAMP-HT participants over usual care patients associated with the incidences of cardiovascular diseases (CVD), end-stage renal disease (ESRD), all-cause mortality and all composite event in selected subgroups by multivariable Cox proportional hazards regressions. (HRs were adjusted for all covariates at baseline.)

**Fig 2b.**
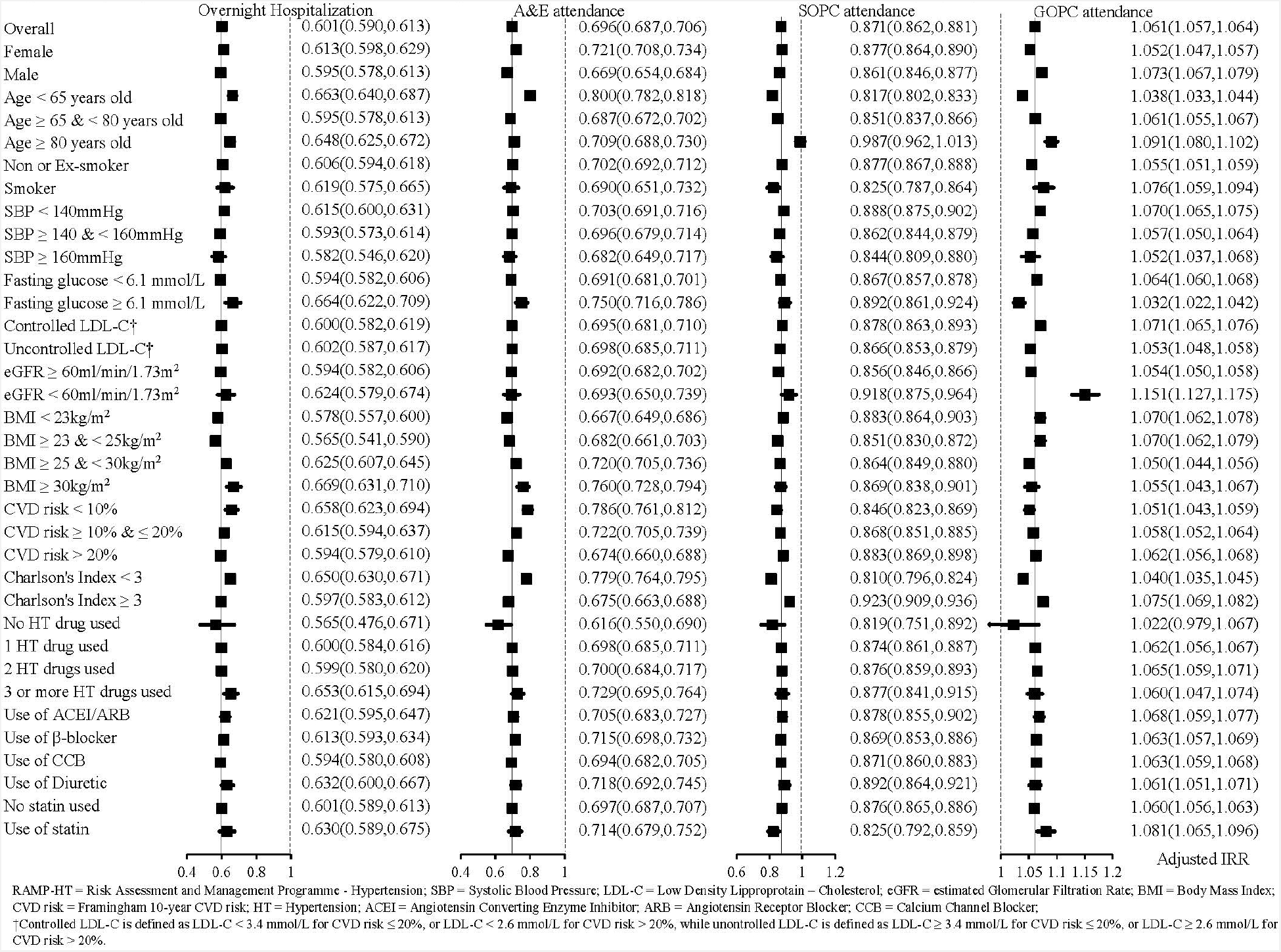
Adjusted incident rate ratios (IRRs) of RAMP-HT participants over usual care patients associated with the number of hospitalization, accident & emergency (A&E) attendance, special outpatient clinic (SOPC) and general outpatient clinic (GOPC) attendance in selected subgroups by negative binomial regressions. (IRRs were adjusted for all covariates at baseline and the corresponding frequency of episode event within one year before baseline.)

## Discussion

Five years after the territory-wide implementation of RAMP-HT – a multi-component, protocol-driven, team-based total CVD risk management programme to augment usual care for primary care patients with hypertension, significant reduction was observed in the absolute risks of incident CVD (−8.0%), ESRD (−1.6%) and all-cause mortality (−10.0%) among RAMP-HT participants compared to patients receiving usual care. The reduction in hypertension-related complications was observed with a decrease in hospital-based service utilization.

The beneficial effects of RAMP-HT on hypertension-related complications and mortality reduction may in part be attributed to the synergistic effects of multiple evidence-based strategies employed to overcome doctors’ therapeutic inertia in optimizing BP and LDL-C control. This was reflected by the higher proportion of RAMP-HT participants being prescribed statins after 5 years. Different from most clinical trials, the RAMP-HT adopted the treat-to-target approach for total CVD risk management; only participants with uncontrolled BP or LDL-C were offered medication titration. As a result, significantly greater proportions of RAMP-HT participants achieved BP and LDL-C targets compared to patients receiving usual care, although absolute between-group differences in BP (−2.2/1.0 mm Hg) and LDL-C (−0.1 mmol/L) were only modest.

Despite the relatively small differences between groups in BP changes over 5 years, the absolute risk reductions (1.6-14.5%) for adverse events were substantial. These findings are comparable to the SPLINT trial, where a specialist nurse-led hypertension/dyslipidaemia clinic resulted in significantly lower mortality (OR 0.55, 95% CI 0.32 to 0.92) after 1 year despite similar BP reduction (mean -1.95/0.79 mm Hg) between the groups.[27] While the magnitude of risk reduction in this study was considerably greater than many large-scale trials on intensive drug regimens,[28-30] it was consistent with the findings from the HOPE-3[31] and ASCOT-LLA[32] trials where the use of statins in addition to BP lowering medications reduced risk of CVD events by 40-50% in individuals with hypertension. A meta-analysis of 61 prospective studies further demonstrated an approximately additive effect of BP and total cholesterol reduction on lowering ischemic heart disease mortality,[33] reinforcing the importance of total CVD risk reduction among patients with hypertension to prevent adverse outcomes.

The add-on 12- to 30-monthly nurse-led risk assessment and nurse intervention sessions may further contribute to CVD reduction through empowering participants with knowledge on hypertension complications and their own CVD risk, and motivating them to comply with various evidence-based measures to manage these risk factors including self-blood pressure monitoring, stop smoking, and adhere to dietary modifications, physical activity and medications.[14, 34] The effectiveness of these interventions may be proxied from the greater likelihood for RAMP-HT participants to be non-smokers, maintain optimal FG levels without pharmacological treatment, and achieve all 5 CVD risk targets after 5 years. Incorporation of a strategic risk stratification in RAMP-HT also allows for better allocation of resources that only patients with relevant risk factors would receive additional multi-disciplinary interventions. As a result, the utilization of other RAMP-HT services and additional allied health interventions were minimal.

Multi-component, multi-modal care has repeatedly demonstrated effectiveness in improving BP control in patients with hypertension.[9, 35] Recently, a multi-dimensional prevention programme targeting CVD-risk factors reduced CVD events by 20% (HR 0.80, 95% CI 0.66 to 0.97) in older Swedish adults (n=3713, >65 years) after 6 years compared to propensity-score-matched controls.[36] Consistently, a post-hoc analysis of the landmark Steno-2 trial in patients with diabetes confirmed that the multi-factorial intervention resulted only in small improvements in disease parameters, but remarkable relative risk reduction in CVD (59%) and all-cause mortality (46%) after a mean follow-up of 13 years.[37] Hence, reduction in adverse outcomes is a function of multiple risk factors improvement and cannot be attributed to BP alone. On the other hand, time saved through task-shifting also allowed more time for doctors to recognize and manage other co-morbidities, complex or urgent issues early, which may contribute to the lower incidence of non-CVD mortality in RAMP-HT participants.

This study describes the actual real-world effectiveness of team-based, multi-component care on a large representative cohort of primary care patients with hypertension over the long-term. Without intensive follow-ups and outcome assessments, our observations reflected how such a model of care benefitted all groups of patients with different levels of literacy, motivation and baseline health in the real-world situation. Several sophisticated analytical techniques including multiple imputations for handling missing data, adjustments for baseline characteristics and fine stratification weightings were used to minimize imbalances in patient baseline characteristics between the comparison groups. Inclusion of an extensive number of confounding variables, sensitivity and subgroups analyses had strengthened the reliability and robustness of the results. The evaluation of the effectiveness of RAMP-HT was comprehensive, including not only surrogate markers but actual CVD events and service utilization. All information obtained from the administrative database were widely used in all clinics and hospitals and were systematically and conscientiously managed by the HA, thus ensuring data accuracy and reliability.

## Limitations

There were also several limitations to this study. First, the prospective cohort design does not prove cause-and-effect. While an array of statistical analyses had been employed to minimize the potential for bias including reverse causality, these biases could not be eliminated entirely. Second, common to studies based on electronic medical records, lifestyle and behavioural factors such as dietary intake and physical activity levels have not been captured in this study. RAMP-HT participants were recruited on a voluntary basis; hence they may inherently be more motivated and adherent to mediation or lifestyle modifications, contributing to possible selection bias, and the potential for the results to be biased away from the null. Third, it is similarly possible that changes in frontline doctors’ practices at clinics offering RAMP-HT has created a globally better practice exposure, offering higher quality of care across all services. While this may be partially mitigated by statistically adjusting for the effects of the care provider clustered within individual clinics, information for the care provider or the clinic was not available to us given concerns for the possibility of reidentifying the patients, and therefore remains a limitation of this study. Fourth, the outcome events relied on the diagnosis code (ICPC-2 and ICD-9-CM) which may be susceptible to misclassification bias. No validation was performed as part of this study to assess the accuracy and completeness of coding. Nevertheless, a previous study showed that only 1.5% and 5.5% of records were miscoded or lacked coding, respectively, for the diagnosis of DM using ICPC-2 in the HA Clinical Management System.[38] Fifth, outcomes measured in this study were limited to those available within the public healthcare system. Service utilization were not recorded if patients received primary care outside the public system. However, patients with chronic diseases and serious complications, e.g., myocardial infarction or stroke, were mostly treated in the heavily-subsidized public system; hence our data should have captured the majority of CVD and ESRD outcomes of patients with hypertension. Lastly, the individual contribution of each element of the multifaceted RAMP-HT programme to its overall effectiveness was not assessed. The RAMP-HT was also conducted in an integrated primary care delivery system fully funded by the government. Direct translation of these findings may be limited to contexts with similar incentives, financial investment and infrastructure support. The ultimate reductions in CVD and mortality observed in this study hence represent a function of change in practices on multiple levels. This reinforces the need to move beyond measurable health outcomes, and to consider the value of non-health outcomes as a determinant for the programme’s success.

## Conclusion

The RAMP-HT – a complex protocol-driven multi-disciplinary intervention integrated into usual primary care – was highly effective in reducing HT-related complications and all-cause mortality in patients with hypertension in the real-world public primary care setting after 5 years. The significant reduction in hospital-based service utilization highlighted its potential to alleviate the public healthcare burden. Further study should determine the cost-effectiveness of RAMP-HT for long-term sustainability of the programme.

## Data Availability

All relevant data are within the manuscript and its Supporting Information files.

## Contributors

EYTY led the conceptualization, study design, writing, and contributed to the methodology. EYFW led the statistical analysis, interpretation, and contributed to the conceptualization, and methodology. ILM contributed to data interpretation and writing. DVKC, WWKK, ML, YCL, JL, LW, MMYW, RLPK, DYTF contributed to methodology, project administration and resources. AKCC contributed to data analysis and interpretation, writing, and administration. CLKL supervised and contributed to the conceptualization, study design, methodology, funding acquisition, data analyses and interpretation. All authors contributed to editing the manuscript. The corresponding author attests that all listed authors meet authorship criteria and that no others meeting the criteria have been omitted. EYTY, EYFW and CLKL are guarantors.

## Data availability statement

As this study is based on data from the Hong Kong Hospital Authority, the authors are not allowed to share data in raw form.

## Supporting Information

S1 Fig. Risk Assessment & Management Programme – Hypertension (RAMP HT) workflow

S2 Fig. Flow chart for patients matching

S1 Table. Definition of the event outcome measures

S2 Table. Data completion rates of RAMP-HT participants and usual care patients at baseline and 5-year follow-up

S3 Table. Baseline characteristics between RAMP-HT participants and usual care patients before and after multiple imputation

S4 Table. Patients’ characteristics between RAMP-HT participants and usual care patients at 5-year follow-up

S5 Table. Sensitivity analyses on effectiveness of all outcome event, diabetes mellitus, cardiovascular disease, end stage renal disease and all-cause mortality between RAMP-HT participants and usual care patients

S6 Table. Sensitivity analyses on comparisons of service utilizations between RAMP-HT participants and usual care patients

## SUPPLEMENTARY FIGURES

**S1 Fig.**
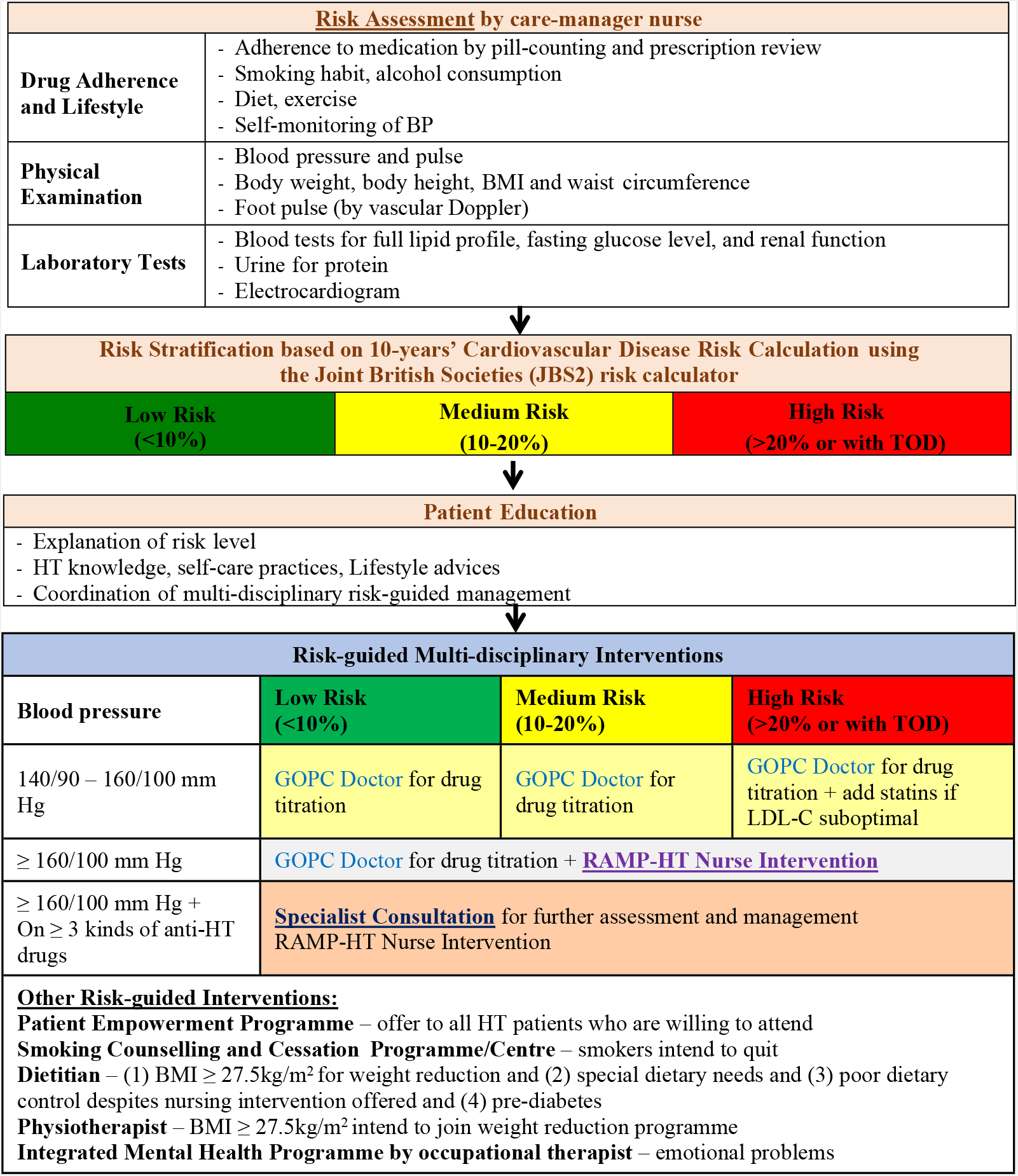
Risk Assessment & Management Programme – Hypertension (RAMP HT) workflow.

**S2 Fig.**
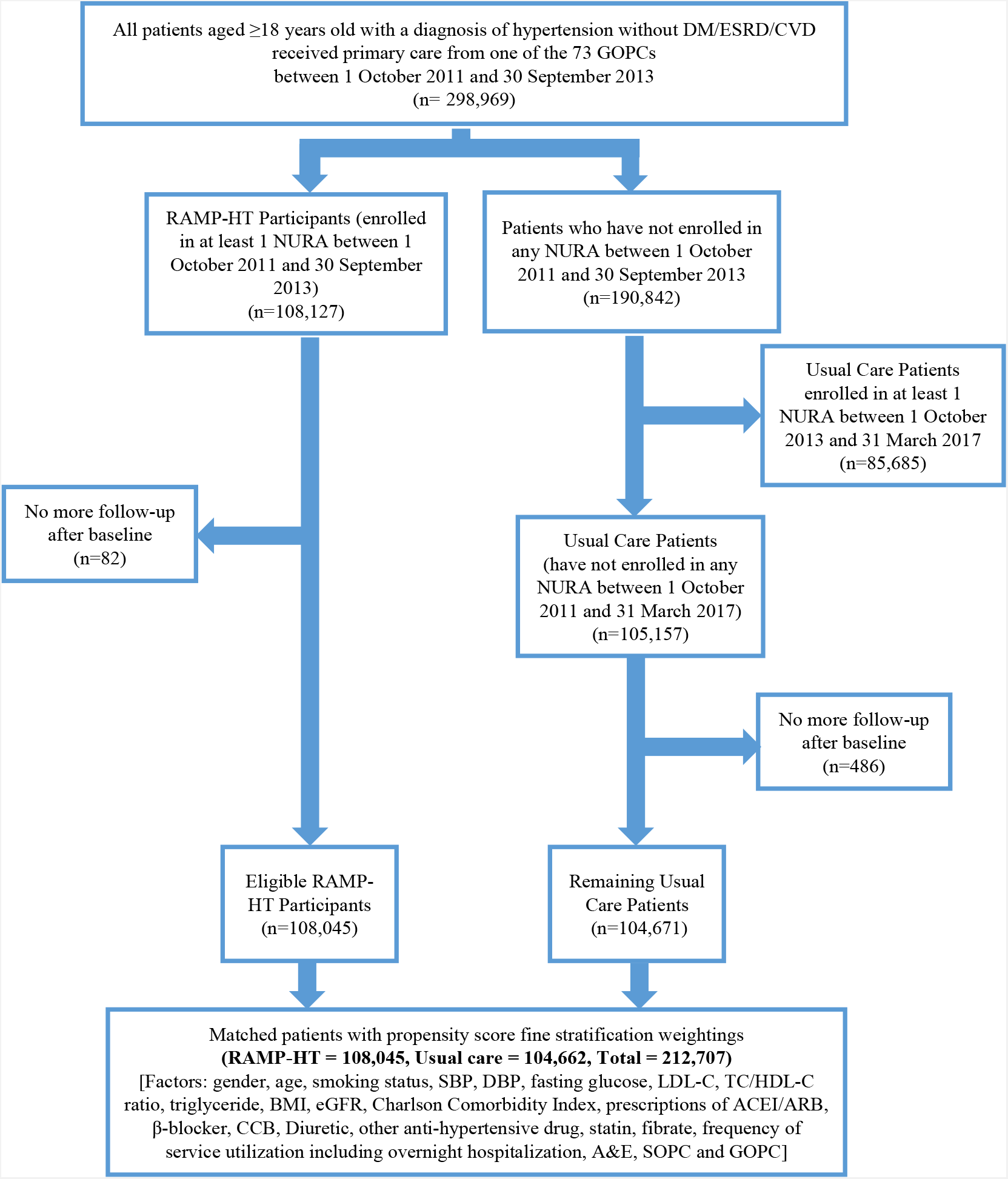
Flow chart for patients matching.

## SUPPLEMENTARY TABLES

**S1 Table.**
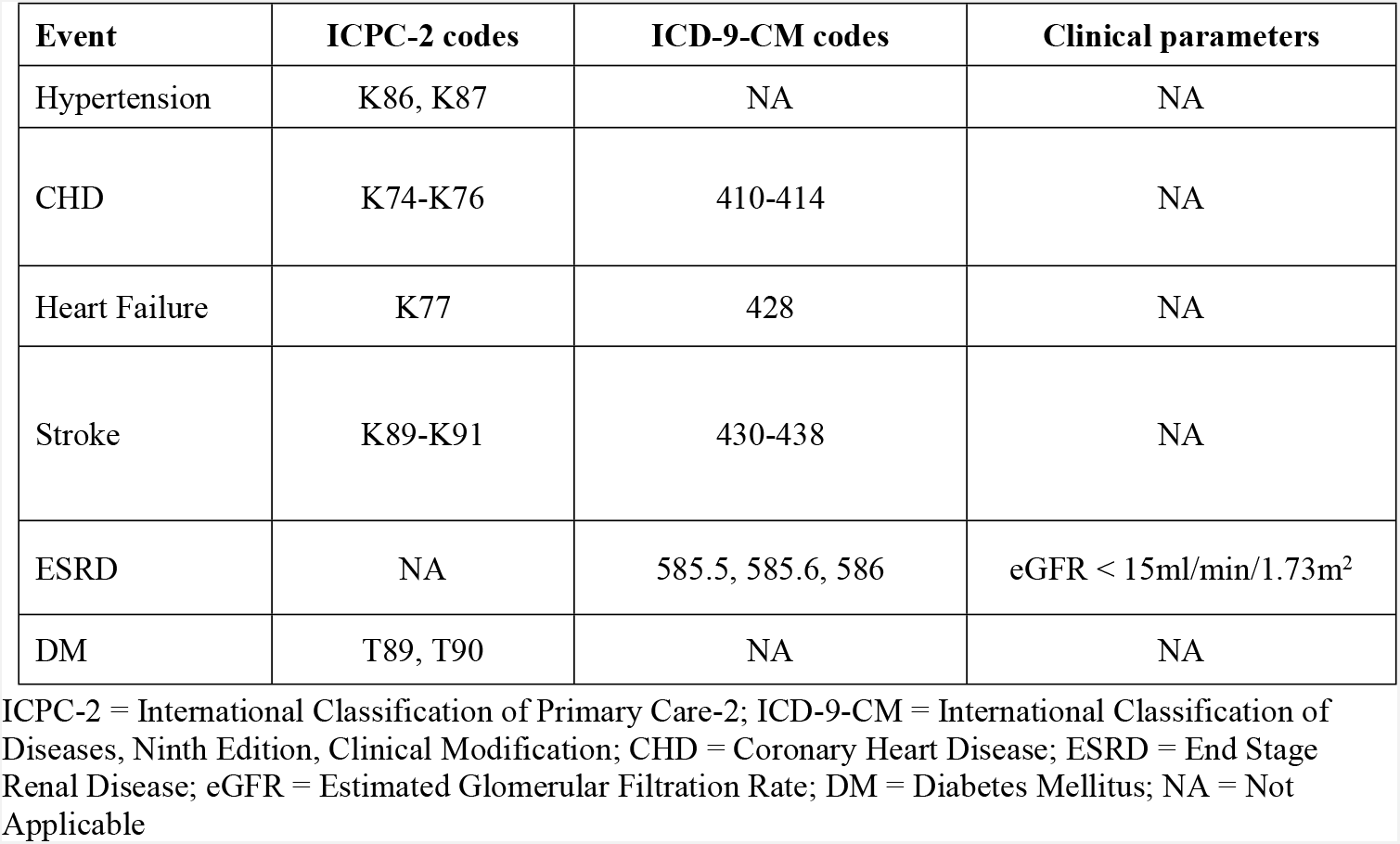
Definition of the event outcome measures.

**S2 Table.** Data completion rates of RAMP-HT participants and usual care patients at baseline and 5-year follow-up.

| Time frame | At baseline |  | At 5 years |  |
| --- | --- | --- | --- | --- |
| Factor | RAMP-HT<br>Participants<br>(N = 108,045) | Usual Care Patients<br>(N = 104,662) | RAMP-HT<br>Participants<br>(N = 108,045) | Usual Care Patients<br>(N = 104,662) |
| **Socio-Demographic (% , n)** |  |  |  |  |
| Gender | 100.00 % (108,045) | 100.00 % (104,662) | 100.00 % (108,045) | 100.00 % (104,662) |
| Age | 100.00 % (108,045) | 100.00 % (104,662) | 100.00 % (108,045) | 100.00 % (104,662) |
| Smoking Status | 99.98 % (108,022) | 98.94 % (103,554) | 99.98 % (108,022) | 98.94 % (103,554) |
| **Clinical Characteristics (% , n)** |  |  |  |  |
| SBP | 100.00 % (108,045) | 99.67 % (104,321) | 92.70 % (100,155) | 80.59 % (84,352) |
| DBP | 100.00 % (108,045) | 99.67 % (104,321) | 92.70 % (100,155) | 80.59 % (84,344) |
| Fasting Glucose | 99.76 % (107,787) | 85.93 % (89,943) | 89.46 % (96,653) | 75.61 % (79,130) |
| LDL-C | 99.63 % (107,647) | 83.79 % (87,696) | 90.61 % (97,900) | 76.06 % (79,602) |
| TC/HDL-C Ratio | 99.86 % (107,891) | 84.12 % (88,044) | 90.73 % (98,026) | 76.15 % (79,695) |
| Triglyceride | 99.88 % (107,917) | 84.63 % (88,577) | 90.74 % (98,035) | 76.22 % (79,778) |
| BMI | 99.79 % (107,816) | 69.92 % (73,181) | 78.58 % (84,900) | 57.08 % (59,740) |
| eGFR | 99.92 % (107,961) | 94.66 % (99,077) | 93.79 % (101,340) | 85.14 % (89,104) |
| Charlson Comorbidity Index | 100.00 % (108,045) | 100.00 % (104,662) | 100.00 % (108,045) | 100.00 % (104,662) |
| Use of ACEI/ARB | 100.00 % (108,045) | 100.00 % (104,662) | 100.00 % (108,045) | 100.00 % (104,662) |
| Use of β-blocker | 100.00 % (108,045) | 100.00 % (104,662) | 100.00 % (108,045) | 100.00 % (104,662) |
| Use of CCB | 100.00 % (108,045) | 100.00 % (104,662) | 100.00 % (108,045) | 100.00 % (104,662) |
| Use of Diuretic | 100.00 % (108,045) | 100.00 % (104,662) | 100.00 % (108,045) | 100.00 % (104,662) |
| Use of other anti-hypertensive drugs | 100.00 % (108,045) | 100.00 % (104,662) | 100.00 % (108,045) | 100.00 % (104,662) |
| Use of Statin | 100.00 % (108,045) | 100.00 % (104,662) | 100.00 % (108,045) | 100.00 % (104,662) |
| Use of Fibrate | 100.00 % (108,045) | 100.00 % (104,662) | 100.00 % (108,045) | 100.00 % (104,662) |
| **Frequency of Service Utilization†** |  |  |  |  |
| Overnight Hospitalization | 100.00 % (108,045) | 100.00 % (104,662) | 100.00 % (108,045) | 100.00 % (104,662) |
| Accident & Emergency | 100.00 % (108,045) | 100.00 % (104,662) | 100.00 % (108,045) | 100.00 % (104,662) |
| Specialist Out-patient Clinic | 100.00 % (108,045) | 100.00 % (104,662) | 100.00 % (108,045) | 100.00 % (104,662) |
| General Out-patient Clinic | 100.00 % (108,045) | 100.00 % (104,662) | 100.00 % (108,045) | 100.00 % (104,662) |

RAMP-HT = Risk Assessment and Management Programme - Hypertension; SBP = Systolic Blood Pressure; DBP = Diastolic Blood Pressure; LDL-C = Low Density Lipoprotein - Cholesterol; TC = Total Cholesterol; HDL-C = High Density Lipoprotein - Cholesterol; BMI = Body Mass Index; eGFR = Estimated Glomerular Filtration Rate; ACEI = Angiotensin Converting Enzyme Inhibitor; ARB = Angiotensin Receptor Blocker; CCB = Calcium Channel Blocker
Notes:
†Service utilization was measured 1) at baseline: from a year before baseline to baseline; 2) at 5 years: from baseline to last follow-up.

**S3 Table.** Baseline characteristics between RAMP-HT participants and usual care patients before and after multiple imputation.

| Factor | Before multiple imputation |  | After multiple imputation |  |
| --- | --- | --- | --- | --- |
|  | RAMP-HT Participants (N = 108,045) | Usual Care Patients (N = 104,662) | RAMP-HT Participants (N = 108,045) | Usual Care Patients (N = 104,662) |
| **Demographics (% , n)** |  |  |  |  |
| Gender |  |  |  |  |
| Female | 59.3 % (64,050) | 56.8 % (59,466) | 59.3 % (64,050) | 56.8 % (59,466) |
| Male | 40.7 % (43,995) | 43.2 % (45,205) | 40.7 % (43,995) | 43.2 % (45,205) |
| Age (mean±SD), year | 65.1±10.8 (108,045) | 68.1±13.5 (104,671) | 65.1±10.8 (108,045) | 68.1±13.5 (104,671) |
| Smoking Status |  |  |  |  |
| Non-Smoker | 93.4 % (100,876) | 91.4 % (94,616) | 93.4 % (100,897) | 91.4 % (95,645) |
| Smoker | 6.6 % (7,146) | 8.6 % (8,938) | 6.6 % (7,148) | 8.6 % (9,026) |
| **Clinical Characteristics (mean±SD)** |  |  |  |  |
| SBP, mm Hg | 136.9±15.5 (108,045) | 136.3±17.3 (104,321) | 136.9±15.5 (108,045) | 136.3±17.3 (104,671) |
| DBP, mm Hg | 76.7±10.6 (108,045) | 75.7±11.3 (104,321) | 76.7±10.6 (108,045) | 75.7±11.3 (104,671) |
| Fasting Glucose, mmol/L | 5.3±0.6 (107,787) | 5.4±0.8 (89,943) | 5.3±0.6 (108,045) | 5.4±0.8 (104,671) |
| LDL-C, mmol/L | 3.2±0.8 (107,647) | 3.2±0.8 (87,696) | 3.2±0.8 (108,045) | 3.2±0.9 (104,671) |
| TC/HDL-C Ratio | 3.9±1.2 (107,891) | 4.0±2.7 (88,046) | 3.9±1.2 (108,045) | 4.0±2.9 (104,671) |
| Triglyceride, mmol/L | 1.4±0.9 (107,917) | 1.5±0.9 (88,577) | 1.4±0.9 (108,045) | 1.5±1.0 (104,671) |
| BMI, kg/m<sup>2</sup> | 25.6±3.9 (107,816) | 25.3±4.1 (73,181) | 25.6±3.9 (108,045) | 25.2±5.2 (104,671) |
| eGFR (% , n) |  |  |  |  |
| ≥ 60ml/min/1.73m<sup>2</sup> | 97.1 % (104,870) | 93.5 % (92,615) | 97.1 % (104,948) | 93.5 % (97,867) |
| < 60ml/min/1.73m<sup>2</sup> | 2.9 % (3,091) | 6.5 % (6,462) | 2.9 % (3,097) | 6.5 % (6,804) |
| Charlson Comorbidity Index | 3.0±1.2 (108,045) | 3.2±1.3 (104,671) | 3.0±1.2 (108,045) | 3.2±1.3 (104,671) |
| Number of HT drugs used (% , n) |  |  |  |  |
| 0 | 1.1 % (1,166) | 3.9 % (4,103) | 1.1 % (1,166) | 3.9 % (4,103) |
| 1 | 53.5 % (57,819) | 53.9 % (56,446) | 53.5 % (57,819) | 53.9 % (56,446) |
| 2 | 37.0 % (40,011) | 34.4 % (36,005) | 37.0 % (40,011) | 34.4 % (36,005) |
| ≥3 | 8.4 % (9,049) | 7.8 % (8,117) | 8.4 % (9,049) | 7.8 % (8,117) |
| Use of ACEI/ARB (% , n) | 20.8 % (22,480) | 18.2 % (19,013) | 20.8 % (22,480) | 18.2 % (19,013) |
| Use of β-blocker (% , n) | 38.5 % (41,632) | 35.4 % (37,063) | 38.5 % (41,632) | 35.4 % (37,063) |
| Use of CCB (% , n) | 72.4 % (78,260) | 68.5 % (71,649) | 72.4 % (78,260) | 68.5 % (71,649) |
| Use of Diuretic (% , n) | 11.8 % (12,762) | 13.1 % (13,679) | 11.8 % (12,762) | 13.1 % (13,679) |
| Use of other anti-hypertensive drugs (% , n) | 10.0 % (10,785) | 11.7 % (12,250) | 10.0 % (10,785) | 11.7 % (12,250) |
| Use of Statin (% , n) | 8.8 % (9,460) | 6.4 % (6,651) | 8.8 % (9,460) | 6.4 % (6,651) |
| Use of Fibrate (% , n) | 1.9 % (2,060) | 1.6 % (1,648) | 1.9 % (2,060) | 1.6 % (1,648) |
| **Frequency of Service Utilization† (mean±SD)** |  |  |  |  |
| Overnight Hospitalization | 0.1±0.5 (108,045) | 0.2±0.8 (104,671) | 0.1±0.5 (108,045) | 0.2±0.8 (104,671) |
| Accident & Emergency | 0.3±0.9 (108,045) | 0.5±1.5 (104,671) | 0.3±0.9 (108,045) | 0.5±1.5 (104,671) |
| Specialist Out-patient Clinic | 1.5±2.7 (108,045) | 1.8±3.2 (104,671) | 1.5±2.7 (108,045) | 1.8±3.2 (104,671) |
| General Out-patient Clinic | 5.5±2.3 (108,045) | 5.8±3.1 (104,671) | 5.5±2.3 (108,045) | 5.8±3.1 (104,671) |

RAMP-HT = Risk Assessment and Management Programme - Hypertension; SBP = Systolic Blood Pressure; DBP = Diastolic Blood Pressure; LDL-C = Low Density Lipoprotein - Cholesterol; TC = Total Cholesterol; HDL-C = High Density Lipoprotein - Cholesterol; BMI = Body Mass Index; eGFR = Estimated Glomerular Filtration Rate; ACEI = Angiotensin Converting Enzyme Inhibitor; ARB = Angiotensin Receptor Blocker; CCB = Calcium Channel Blocker; SD = Standard Deviation
Notes:
† Service utilization was measured from a year before baseline to baseline

**S4 Table.**
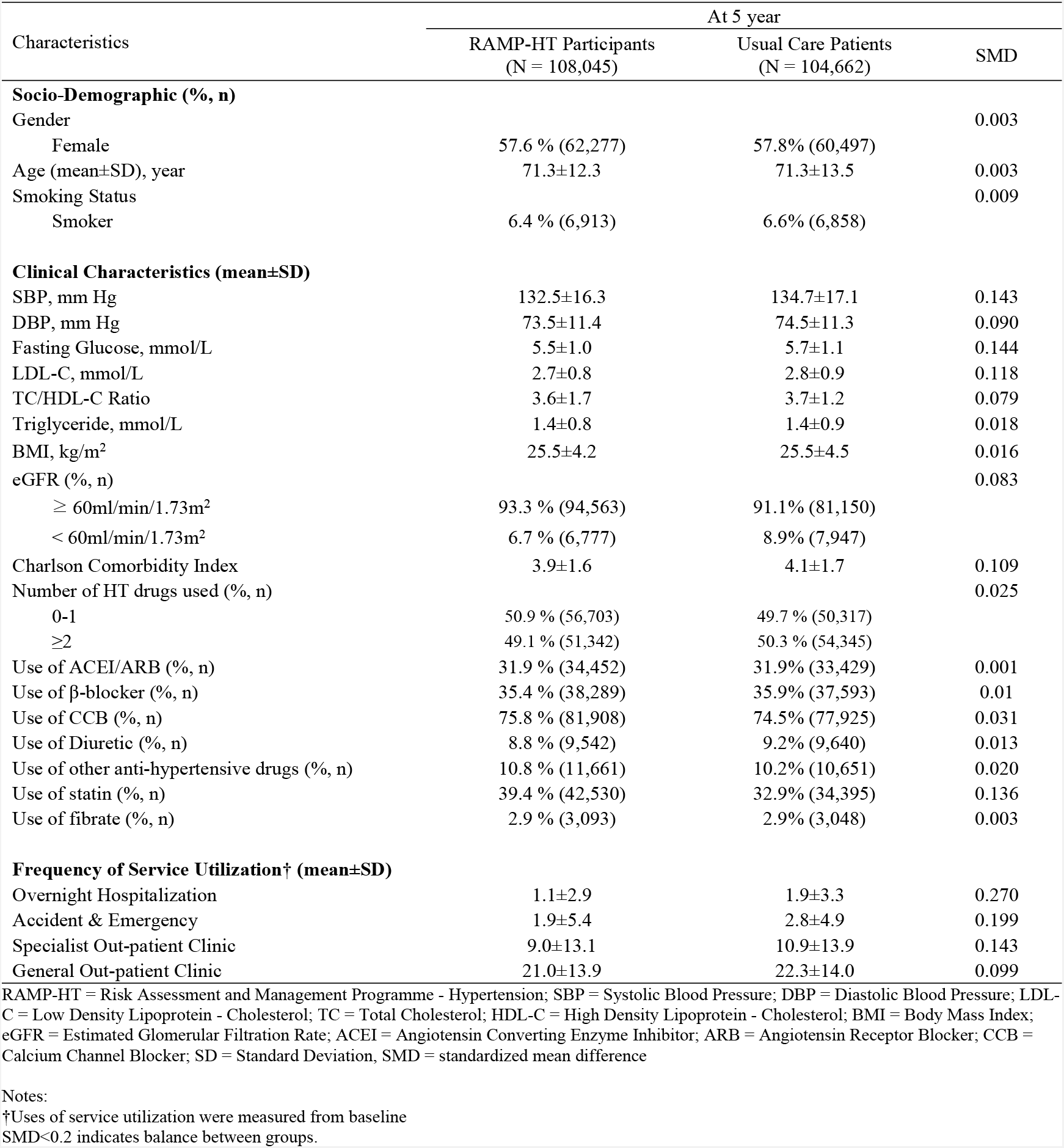
Patients’ characteristics between RAMP-HT participants and usual care patients at 5-year follow-up.

| Characteristics | At 5 year |  |  |
| --- | --- | --- | --- |
|  | RAMP-HT Participants<br>(N = 108,045) | Usual Care Patients<br>(N = 104,662) | SMD |
| **Socio-Demographic (% , n)** |  |  |  |
| Gender |  |  | 0.003 |
| Female | 57.6 % (62,277) | 57.8% (60,497) |  |
| Age (mean±SD), year | 71.3±12.3 | 71.3±13.5 | 0.003 |
| Smoking Status |  |  | 0.009 |
| Smoker | 6.4 % (6,913) | 6.6% (6,858) |  |
| **Clinical Characteristics (mean±SD)** |  |  |  |
| SBP, mm Hg | 132.5±16.3 | 134.7±17.1 | 0.143 |
| DBP, mm Hg | 73.5±11.4 | 74.5±11.3 | 0.090 |
| Fasting Glucose, mmol/L | 5.5±1.0 | 5.7±1.1 | 0.144 |
| LDL-C, mmol/L | 2.7±0.8 | 2.8±0.9 | 0.118 |
| TC/HDL-C Ratio | 3.6±1.7 | 3.7±1.2 | 0.079 |
| Triglyceride, mmol/L | 1.4±0.8 | 1.4±0.9 | 0.018 |
| BMI, kg/m<sup>2</sup> | 25.5±4.2 | 25.5±4.5 | 0.016 |
| eGFR (% , n) |  |  | 0.083 |
| ≥ 60ml/min/1.73m<sup>2</sup> | 93.3 % (94,563) | 91.1% (81,150) |  |
| < 60ml/min/1.73m<sup>2</sup> | 6.7 % (6,777) | 8.9% (7,947) |  |
| Charlson Comorbidity Index | 3.9±1.6 | 4.1±1.7 | 0.109 |
| Number of HT drugs used (% , n) |  |  | 0.025 |
| 0-1 | 50.9 % (56,703) | 49.7 % (50,317) |  |
| ≥2 | 49.1 % (51,342) | 50.3 % (54,345) |  |
| Use of ACEI/ARB (% , n) | 31.9 % (34,452) | 31.9% (33,429) | 0.001 |
| Use of β-blocker (% , n) | 35.4 % (38,289) | 35.9% (37,593) | 0.01 |
| Use of CCB (% , n) | 75.8 % (81,908) | 74.5% (77,925) | 0.031 |
| Use of Diuretic (% , n) | 8.8 % (9,542) | 9.2% (9,640) | 0.013 |
| Use of other anti-hypertensive drugs (% , n) | 10.8 % (11,661) | 10.2% (10,651) | 0.020 |
| Use of statin (% , n) | 39.4 % (42,530) | 32.9% (34,395) | 0.136 |
| Use of fibrate (% , n) | 2.9 % (3,093) | 2.9% (3,048) | 0.003 |
| **Frequency of Service Utilization† (mean±SD)** |  |  |  |
| Overnight Hospitalization | 1.1±2.9 | 1.9±3.3 | 0.270 |
| Accident & Emergency | 1.9±5.4 | 2.8±4.9 | 0.199 |
| Specialist Out-patient Clinic | 9.0±13.1 | 10.9±13.9 | 0.143 |
| General Out-patient Clinic | 21.0±13.9 | 22.3±14.0 | 0.099 |

RAMP-HT = Risk Assessment and Management Programme - Hypertension; SBP = Systolic Blood Pressure; DBP = Diastolic Blood Pressure; LDL-C = Low Density Lipoprotein - Cholesterol; TC = Total Cholesterol; HDL-C = High Density Lipoprotein - Cholesterol; BMI = Body Mass Index; eGFR = Estimated Glomerular Filtration Rate; ACEI = Angiotensin Converting Enzyme Inhibitor; ARB = Angiotensin Receptor Blocker; CCB = Calcium Channel Blocker; SD = Standard Deviation, SMD = standardized mean difference
Notes:
†Uses of service utilization were measured from baseline
SMD<0.2 indicates balance between groups.

**S5 Table.** Sensitivity analyses on effectiveness of all outcome event, diabetes mellitus, cardiovascular disease, end stage renal disease and all-cause mortality between RAMP-HT participants and usual care patients.

| Event | Sensitivity analyses |  |  |  |  |  |  |  |  |  |  |  |  |  |  |  |  |  |
| --- | --- | --- | --- | --- | --- | --- | --- | --- | --- | --- | --- | --- | --- | --- | --- | --- | --- | --- |
|  | (1) |  |  | (2) |  |  | (3) |  |  | (4) |  |  | (5) |  |  | (6) |  |  |
|  | ARR | HR† | NNT | ARR | HR† | NNT | ARR | HR† | NNT | ARR | HR† | NNT | ARR | HR† | NNT | ARR | HR† | NNT |
| All outcome event | 12.0% | 0.61* | 10 | 9.5% | 0.59* | 12 | 14.5% | 0.55* | 9 | 12.0% | 0.59* | 12 | 9.2% | 0.60* | 12 | 12.0% | 0.57* | 10 |
| CVD | 6.4% | 0.66* | 23 | 5.8% | 0.62* | 17 | 8.0% | 0.59* | 15 | 7.4% | 0.61* | 17 | 5.8% | 0.62* | 17 | 7.4% | 0.59* | 15 |
| CHD | 1.9% | 0.71* | 65 | 2.0% | 0.66* | 47 | 2.6% | 0.64* | 41 | 2.6% | 0.65* | 46 | 2.1% | 0.66* | 46 | 2.6% | 0.63* | 41 |
| Heart Failure | 2.5% | 0.56* | 62 | 2.1% | 0.54* | 52 | 3.2% | 0.50* | 38 | 2.7% | 0.53* | 53 | 2.2% | 0.53* | 49 | 2.7% | 0.51* | 44 |
| Stroke | 3.2% | 0.67* | 80 | 3.0% | 0.63* | 35 | 4.0% | 0.61* | 30 | 3.7% | 0.63* | 36 | 2.8% | 0.64* | 36 | 3.7% | 0.61* | 32 |
| ESRD | 1.3% | 0.54* | 95 | 0.7% | 0.62* | 155 | 1.6% | 0.50* | 79 | 1.4% | 0.54* | 119 | 0.7% | 0.63* | 158 | 1.3% | 0.52* | 93 |
| DM | 4.3% | 0.78* | 91 | 4.0% | 0.79* | 33 | 4.6% | 0.70* | 23 | 6.3% | 0.76* | 28 | 4.5% | 0.78* | 30 | 6.3% | 0.67* | 19 |
| All-Cause Mortality | 8.2% | 0.55* | 21 | 6.0% | 0.54* | 20 | 10.0% | 0.48* | 14 | 7.4% | 0.55* | 22 | 5.6% | 0.57* | 22 | 7.4% | 0.53* | 18 |
| CVD Mortality | 3.0% | 0.55* | 61 | 2.4% | 0.50* | 47 | 3.9% | 0.46* | 32 | 3.1% | 0.51* | 51 | 2.3% | 0.53* | 50 | 3.1% | 0.49* | 41 |
| Non-CVD Mortality | 5.3% | 0.55* | 62 | 3.5% | 0.56* | 32 | 6.1% | 0.49* | 21 | 4.3% | 0.58* | 59 | 3.3% | 0.59* | 35 | 4.3% | 0.56* | 30 |

(1) = Excluded patients with less than 1-year follow-up; (2) = Multiple imputation with one-to-one propensity score matching; (3) = Multiple imputation without propensity score matching or weightings; (4) = Fine stratification weightings without multiple imputation; (5) = Propensity score matching only without multiple imputation; (6) = Complete case analysis
RAMP-HT = Risk Assessment and Management Programme - Hypertension; CVD = Cardiovascular Disease; CHD = Coronary Heart Disease; ESRD = End Stage Renal Disease; DM = Diabetes Mellitus; ARR = Absolute Risk Reduction; HR = Hazard Ratio; NNT = Number Needed to Treat;
\* P-value <0.05.
† Hazard ratios were adjusted by gender, age, smoking status, SBP, DBP, fasting glucose, LDL-C, TC/HDL-C ratio, triglyceride, BMI, eGFR, Charlson Comorbidity Index and the usages of ACEI/ARB, β-blocker, CCB, Diuretic, other anti-hypertensive drugs, statin and fibrate at baseline by multivariable Cox proportional hazard regression/multivariable Cox proportional hazard regression with fine stratification weighting, as appropriate

**S6 Table.** Sensitivity analyses on comparisons of service utilizations between RAMP-HT participants and usual care patients.

|  | Sensitivity analyses |  |  |  |  |  |  |  |  |  |  |  |
| --- | --- | --- | --- | --- | --- | --- | --- | --- | --- | --- | --- | --- |
|  | (1) |  | (2) |  | (3) |  | (4) |  | (5) |  | (6) |  |
| Frequency of Event during the Period | Rate | IRR† | Rate | IRR† | Rate | IRR† | Rate | IRR† | Rate | IRR† | Rate | IRR† |
| Overnight Hospitalization | -12.4 | 0.62* | -12.6 | 0.61* | -21.7 | 0.57* | -17.0 | 0.62* | -12.1 | 0.62* | -17.2 | 0.59* |
| A&E Attendance | -13.1 | 0.72* | -13.6 | 0.70* | -24.6 | 0.68* | -19.8 | 0.72* | -13.5 | 0.72* | -20.1 | 0.70* |
| SOPC Attendance | -17.3 | 0.88* | -23.5 | 0.85* | -39.7 | 0.85* | -41.2 | 0.85* | -22.5 | 0.85* | -42.6 | 0.83* |
| GOPC Attendance | 14.6 | 1.05* | 0.3 | 1.05* | 9.2 | 1.06* | -5.4 | 1.04* | -1.7 | 1.04* | -8.5 | 1.04* |

(1) = Excluded patients with less than 1-year follow-up; (2) = Multiple imputation with one-to-one propensity score matching; (3) = Multiple imputation without propensity score matching or weightings; (4) = Fine stratification weightings without multiple imputation; (5) = Propensity score matching only without multiple imputation; (6) = Complete case analysis
RAMP-HT = Risk Assessment and Management Programme - Hypertension; A&E = Accident and Emergency; SOPC = Specialist Outpatient Clinic; GOPC = General Outpatient Clinic; IRR = Incidence Rate Ratio
The unit for incidence rate is Cases/100 Person Years.
\* P-value <0.05.
† All incidence rate ratios were adjusted by gender, age, smoking status, SBP, DBP, fasting glucose, LDL-C, TC/HDL-C ratio, triglyceride, BMI, eGFR, Charlson Comorbidity Index and the usages of ACEI/ARB, β-blocker, CCB, Diuretic, other anti-hypertensive drugs, statin and fibrate, and the corresponding number of service utilizations at baseline by multivariable negative binomial regression/multivariable negative binomial regression with fine stratification weighting, as appropriate

